# Associations between solid fuel use and early child development among 3 to 4 years old children in Bangladesh: Evidence from a nationally representative survey

**DOI:** 10.1101/2020.11.12.20230672

**Authors:** Juwel Rana, Patricia Luna Gutierrez, Syed Emdadul Haque, José Ignacio Nazif-Muñoz, Dipak K. Mitra, Youssef Oulhote

## Abstract

**Background:** Household Air Pollution (HAP) from solid fuel use (SFU) may have impacts on children’s health in low-resources countries. Despite these potential health effects, SFU is still highly prevalent in Bangladesh.

**Objectives:** This study was conducted to assess the associations between SFU and early childhood development index (ECDI) among under-five children in Bangladesh and explore the potential effect modification by sex and urbanicity.

**Materials and methods:** This cross-sectional study used Bangladesh Multiple Indicator Cluster Survey (MICS) 2019, a nationally representative survey data collected by UNICEF from all 64 districts in Bangladesh. The ECDI consisted of ten different items across four developmental domains: literacy-numeracy, physical, social-emotional development, and learning skills in the early years of life (36 to 59 months). A total of 9,395 children aged 36 to 59 months were included in this analysis. We used multilevel Poisson regression models with a robust variance where SFU was a proxy indicator for HAP exposure.

**Results:** Children exposed to SFU were 1.47 times more likely to be not developmentally on track (95% CI: 1.25, 1.73; <0.001) compared to children with no SFU exposure. Two sub-domains explained these associations, SFU was significantly associated with socio-emotional development (prevalence ratio [PR]: 1.17; 95% CI: 1.01, 1.36; p=0.035), and learning-cognitive development (PR: 1.90; 95% CI: 1.39, 2.60; p<0.001). Associations between SFU and ECDI were not significantly different (p-difference=0.210) between girls (PR: 1.64; 95% CI: 1.31, 2.07) and boys (PR: 1.37; 95% CI: 1.13, 1.65). Likewise, urbanicity did not modify the associations between SFU and ECDI outcomes.

**Conclusion:** Bangladeshi children aged 36-59 months exposed to SFU exhibited delays in childhood development compared to unexposed children. Public health policies should promote a better early life environment for younger children to meet their developmental milestones by reducing the high burden of HAP exposure in low-resource settings where most disadvantaged kids struggle to reach their full developmental potentials.

## 1. Introduction

Household Air Pollution (HAP) from cooking with solid fuels contributed to at least 3.6 million cases of premature mortality in low- and middle-income countries (LMICs) in 2016(1), making solid fuel use (SFU) a major risk factor for health in LMICs (2). SFU is most prevalent in Africa and Southeast Asia (3). This problem is particularly acute in Bangladesh; 87% of households (2017) rely on biomass fuels and are at risk of SFU induced harmful effects (4). Through its Sustainable Development Goals (SDGs), the United Nations has reaffirmed its commitment to ensure better early child development and protect populations from HAP (5). These new challenges of the SDG era have a transversal connection with other SDGs, including those mapped to improve education, reduce the impact of climate change, lower poverty, and achieve gender equality (5–7).

It is well established that SFU is a major threat to children’s health (8–10). It has been associated with higher risks of respiratory diseases (8,10), neonatal mortality, infant mortality, low birth weight (11), a variety of cancers, and other outcomes (12). Recent animal and human studies also suggest that air pollution can affect brain development (13,14). For instance, exposure to SFU induced particulate matter (PM10, PM2.5,) CO and NO2 are associated with damage to the central nervous system (local systematic inflammation, oxidative stress, neurotoxicity) (15), schizophrenia (16), which increased the risk for developing attention-deficit hyperactive disorder (ADHD) (17), aggression (18), changes in white matter (19) and damage in brain structure, predominantly when the exposure is experienced during early stages of life (including perinatal exposures) (20). Looking at early exposure is critically important because the first years of life constitute a critical window for brain development (20). Moreover, exposure to SFU disproportionally affects women and children, mostly involved in cooking (8,21–23).

However, most of these studies are limited to developed countries (15,20). Recent evidence suggests that about 25 million children under 5 in LMICs are at risk of not reaching their full potential due to poverty, stunting, nutrient deficiency, violence, heavy metal exposure, social, biological, and physiological factors (24,25). Furthermore, early childhood development (ECD) delays have been associated with lags in adult productivity and health challenges later in life (3,25,26). In light of these indications, the study of the potential effects of SFU on child development in LMICs becomes a key area of investigation when addressing child development in the context of SDGs.

Studies assessing HAP and ECD in LMCIs are slowly emerging. Two studies in Ghana (23,27) found a negative association between school attendance and energy poverty and SFU and ECD. While the studies carried out in Ghana move our understanding of SFU with child development and education forward, these studies did not account for seasonality nor cooking place; thus, their estimates could be biased. On the other hand, two studies assessed how environmental exposures such as arsenic and manganese affected children’s intellectual functions in Bangladesh (28,29). Studies in this country, however, did not examine the association between SFU and ECD. To fill in the gaps in this literature, this study aims to assess the association between SFU and ECDI outcomes among under-five children in Bangladesh and explore the potential effect modification by sex and urbanicity in these associations.

## 2. Materials and Methods

### 2.1. Study participants and data resources

We conducted this cross-sectional study using the latest round of Bangladesh Multiple Indicator Cluster Survey (MICS) 2019. The MICS is a nationally representative household survey that generates reliable, statistically sound, and comparable international records (30). This dataset included 100 crucial indicators of the health and well-being condition in which women and children live in LMICs. Data were collected through five different face-to-face questionnaires gathering information about basic demographic indicators of women (aged 15–49 years), children (aged 5-17), and younger children (under five years).

The MICS 2019 includes a representative sample from eight divisions and all 64 districts located in urban and rural areas of Bangladesh in which information of 9,395 children 3-4 years old was collected (31). Detailed information on the survey is available at http://mics.unicef.org/ (30).

### 2.2. Early child development indicators

The Early Childhood Development Index (ECDI) is a development indicator that assesses whether a child is developmentally on track regarding specific milestones. It was first introduced as part of the fourth round of the MICS in the Questionnaire for Children under-5 and provided the first population-based indicator of early childhood development comparable at the international level (6,31). The index consists of ten different items across four developmental domains: literacy-numeracy (3 items), physical (2 items), social-emotional development (3 items), and learning skills (2 items) across the early years of life (36 to 59 months) (31). A child is considered developmentally on track if the child is on track in at least three of the four domains:

#### Literacy-numeracy

A child is considered developmentally on track if they can identify at least ten letters of the alphabet, read at least four simple words, and know the name or recognize the symbols of all numbers from 1 to 10 (31).

#### Physical

A child is considered developmentally on track if they can pick up a small object with two fingers and if the mother or primary caregiver does not indicate that the child is sometimes too sick to play (31).

#### Social-emotional

A child is considered developmentally on track if they can perform two of the following: if the child gets along with other children; if the child does not kick, bite or hit other children; and if the child does not get distracted easily (31).

#### Learning-cognition

A child is considered developmentally on track if they can follow simple directions and do it independently when indicated (31).

### 2.3. Solid Fuel Use

SFU is considered here as a proxy for HAP. A child is considered “exposed” if the household reports the use of coal, charcoal, wood, straw/shrubs, animal dung, or agricultural crop residue as a primary source of energy for cooking in the traditional solid fuel cookstove and is considered “non-exposed” if the household reports the use of electricity, liquefied petroleum gas, natural gas and/or biogas as a primary source of energy for cooking in the clean cookstove.

### 2.4. Covariates

Based on prior literature (10,17,20,23,25,32,33), variables that are known to be associated with early child development, such as mother’s characteristics (age, education, and marital status), children’s characteristics (age, sex, size, iodine supplementation, stunting, attendance in early child development education program (ECDEP), caregiver’s stimulation), spatial characteristics (region, urbanicity-urban & rural, seasonality) and household’s characteristics (paternal education, household wealth quintiles, cooking place, types of cookstoves and chimney/fan-is there any chimney/fan if households have solid fuel cookstove) were included as covariates. While breastfeeding status for children aged 3-5 was unavailable, we use stunting as a proxy for nutritional status. We have reconstructed household wealth quintiles excluding types of cooking fuels used in the households to avoid over adjustment in inferential models as the original household wealth quintiles (index of available household assets) provided by the United Nations Children’s Fund (UNICEF) included types of fuels to construct household wealth index. The household wealth quintiles were categorized into five quintiles, with the lowest quintile reflecting the poorest households and highest quintile the wealthiest households.

### 2.5. Statistical analysis

Descriptive statistics were used to provide general features of the data. The distribution and prevalence of global ECDI and its sub-domains were calculated by each of the covariates.

Odds ratios estimated using logistic regression can significantly overestimate relative risk (RR) when the outcome is common (34). We used a multilevel Poisson regression model with robust variance to address this, including the survey weights to examine the associations between SFU, ECDI, and its four subdomains (10,34). Results from the multilevel Poisson regression model are presented as a prevalence ratio (PRs) with 95% confidence intervals (95% CI); we considered a p-value of 0.05 as the level of statistical significance. Additional analyses explore the associations between SFU and ECDI modified by sex (boys/girls) and urbanicity status (Rural/urban). We used random subject effects accounting for cluster and stratum. Final models were adjusted for the following fixed effects: child sex, child age, maternal age, maternal education, household wealth quintiles, iodine status, stunting, attendance to ECDEP, cooking place, season and urbanicity. Differences between estimates for boys and girls and urban and rural areas were estimated using a p-difference extracted using a Wald test for the interaction term. All analyses were conducted in R statistical software version 4.2 (35) and Stata version 16 (36).

### 2.6 Data and Code Availability

All data files are available from the MICS program database: https://mics.unicef.org/surveys. Codes are available upon request.

## 3. Results

### 3.1. Prevalence of ECD outcomes and associated characteristics

The prevalence of ECD outcomes, characteristics of the study children aged 3-4 years, their parental and household information are presented in Table 1. A total of 9,395 children aged 3-4 years were included in this analysis. The prevalence of children not being developmentally on track was 25.26% in overall ECDI. For the four subdomains: 71.14% of children were not developmentally on track in literacy-numeracy, 27.20% in socio-emotional development, 8.60% in learning, and 1.38% in physical development (**Table 1**). On the other hand, the prevalence of SFU as a primary source of energy for cooking and use of traditional solid fuel cookstoves was 81.37% in Bangladeshi households.

**Table 1.**
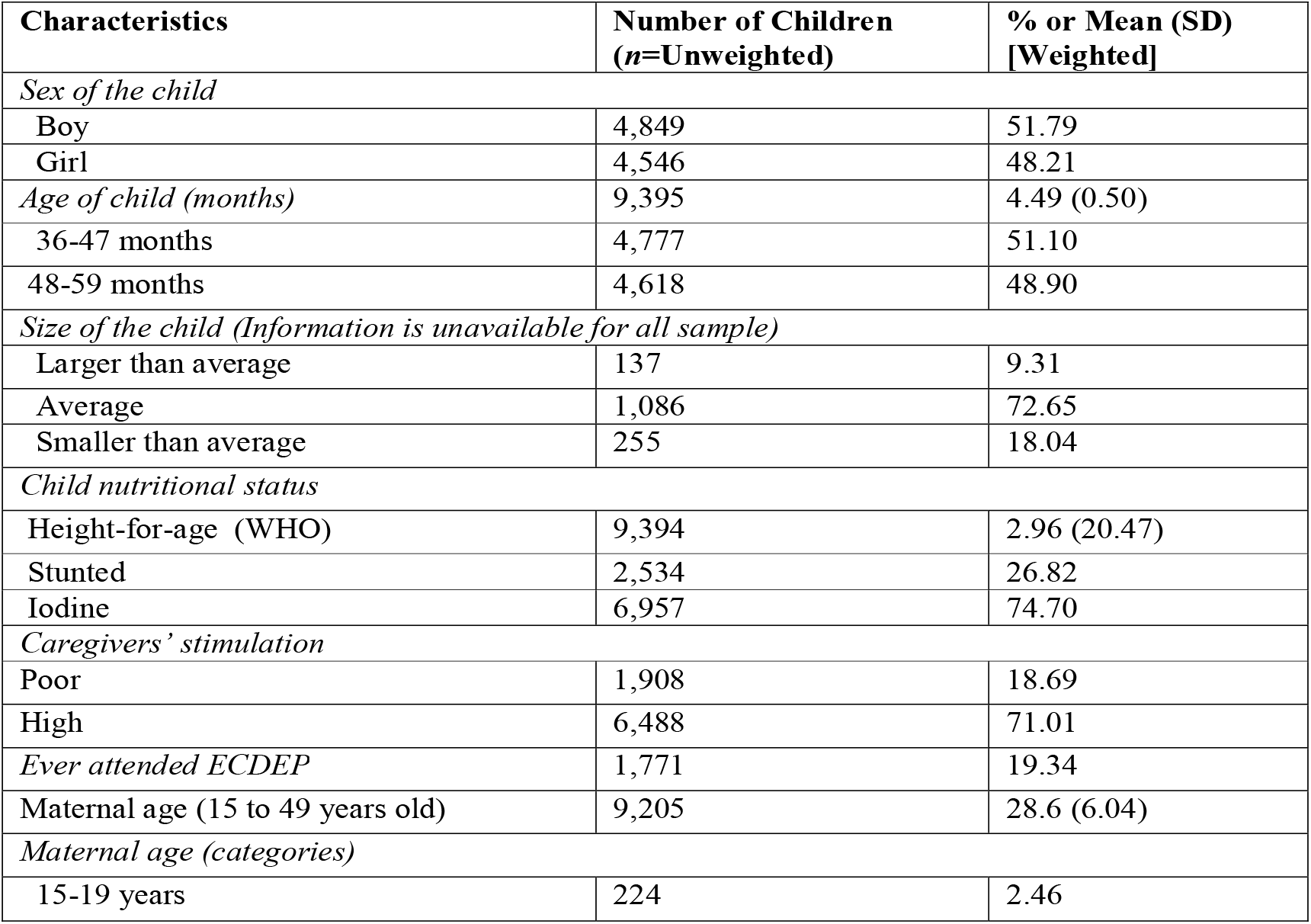
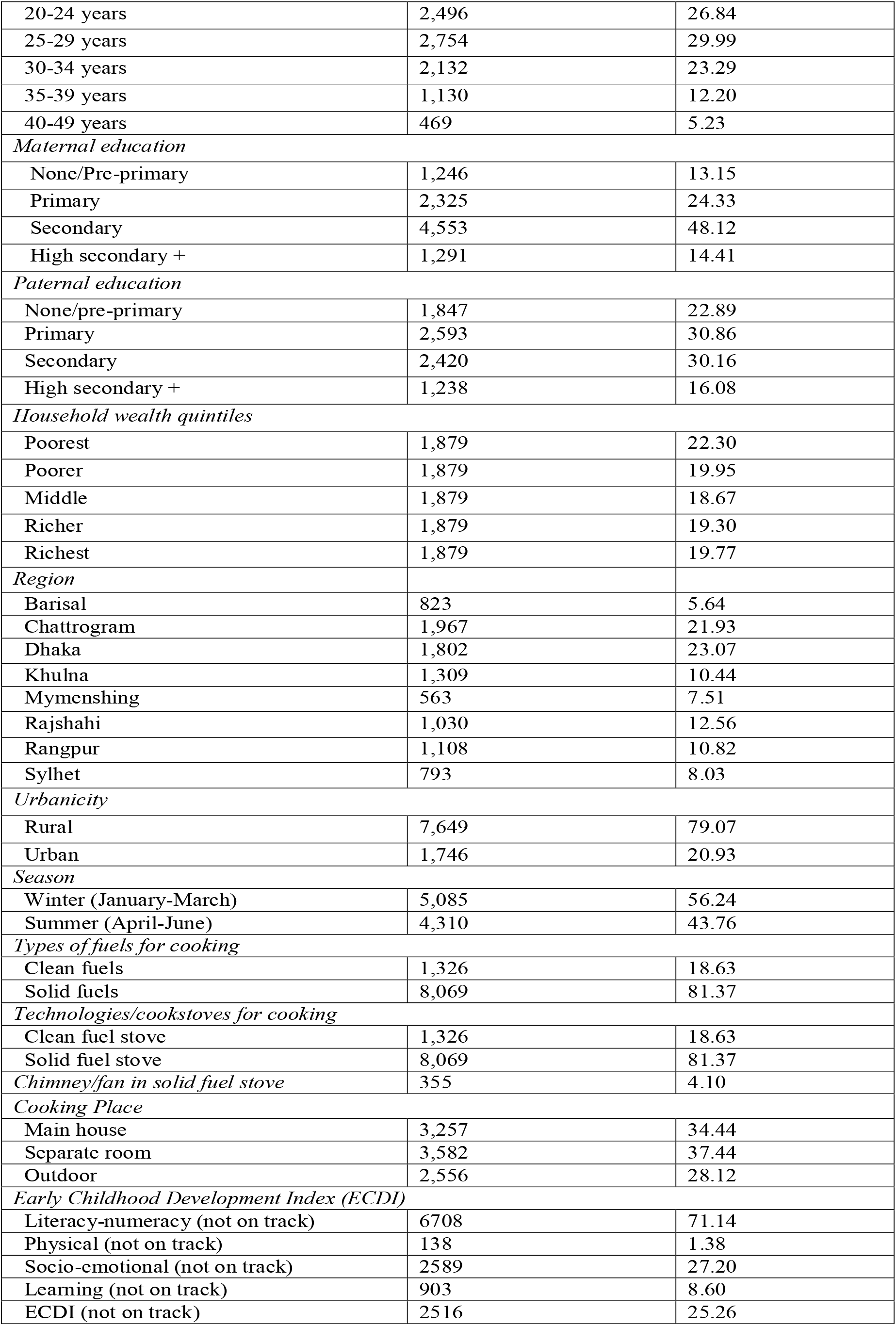
Sample characteristics of 36 to 59 months old children in Bangladesh (Weighted Percentage or mean and n=Unweighted)

Overall, 51.79% of the children were boys, 51.10% were between 36 and 47 months of age, 19.34% attended an ECDEP, 71.01% received sufficient support from caregivers, 74.70% received intake of iodine, 26.82% were stunted, and 79.07% lived in rural settings. Mothers’ characteristics indicate that 56.83% of mothers were between 20 and 29 years old, and 37.48% had less than or equal to primary education. About 34.44% of households have their cooking place in the main house, 81.37% of households used solid fuel cookstoves, and only 4.10% of them are equipped with a chimney or a fan.

The results from univariate associations of ECD outcomes by each sub-domain with sample characteristics are presented in **Table 2**.

**Table 2.**
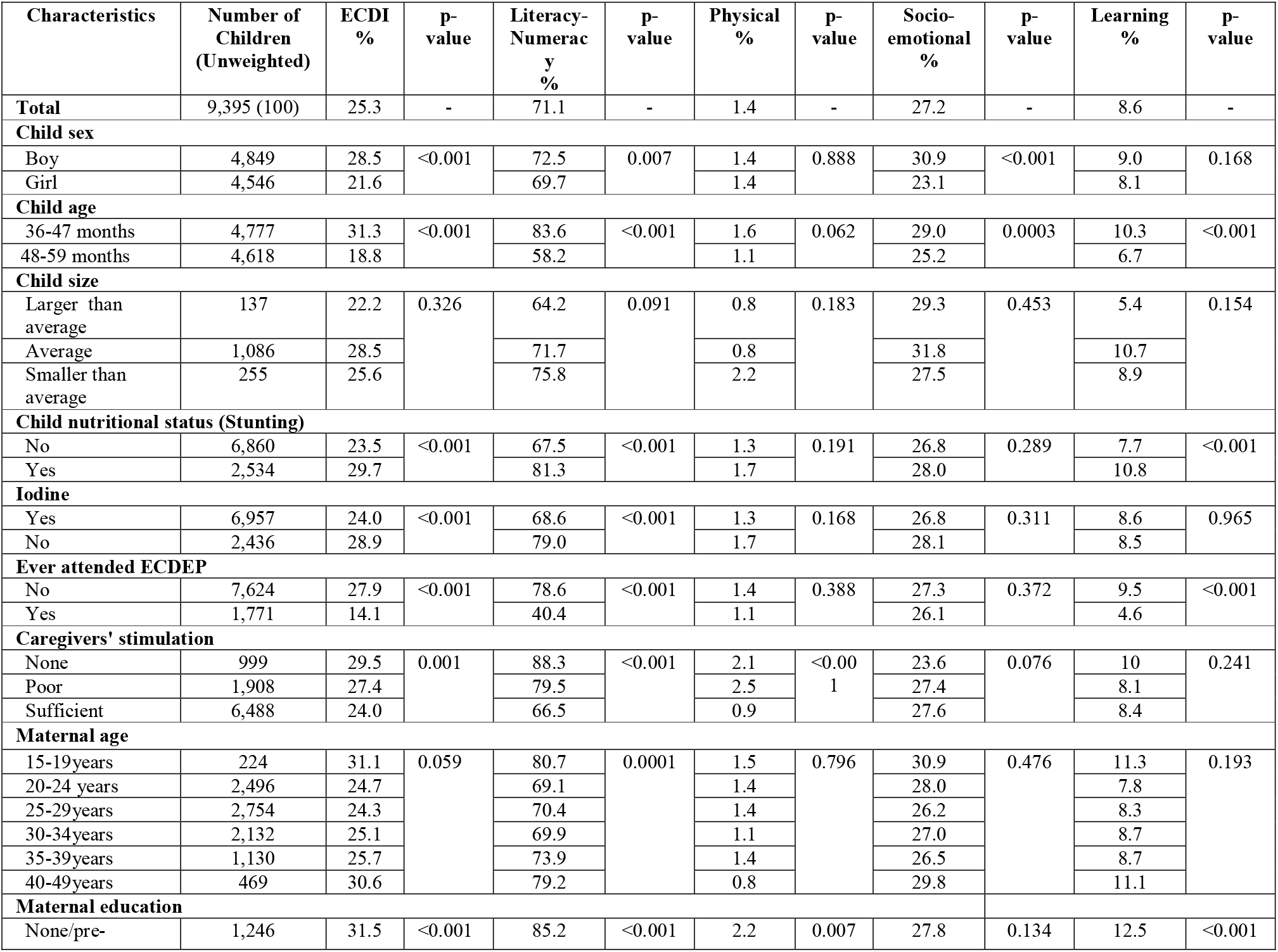
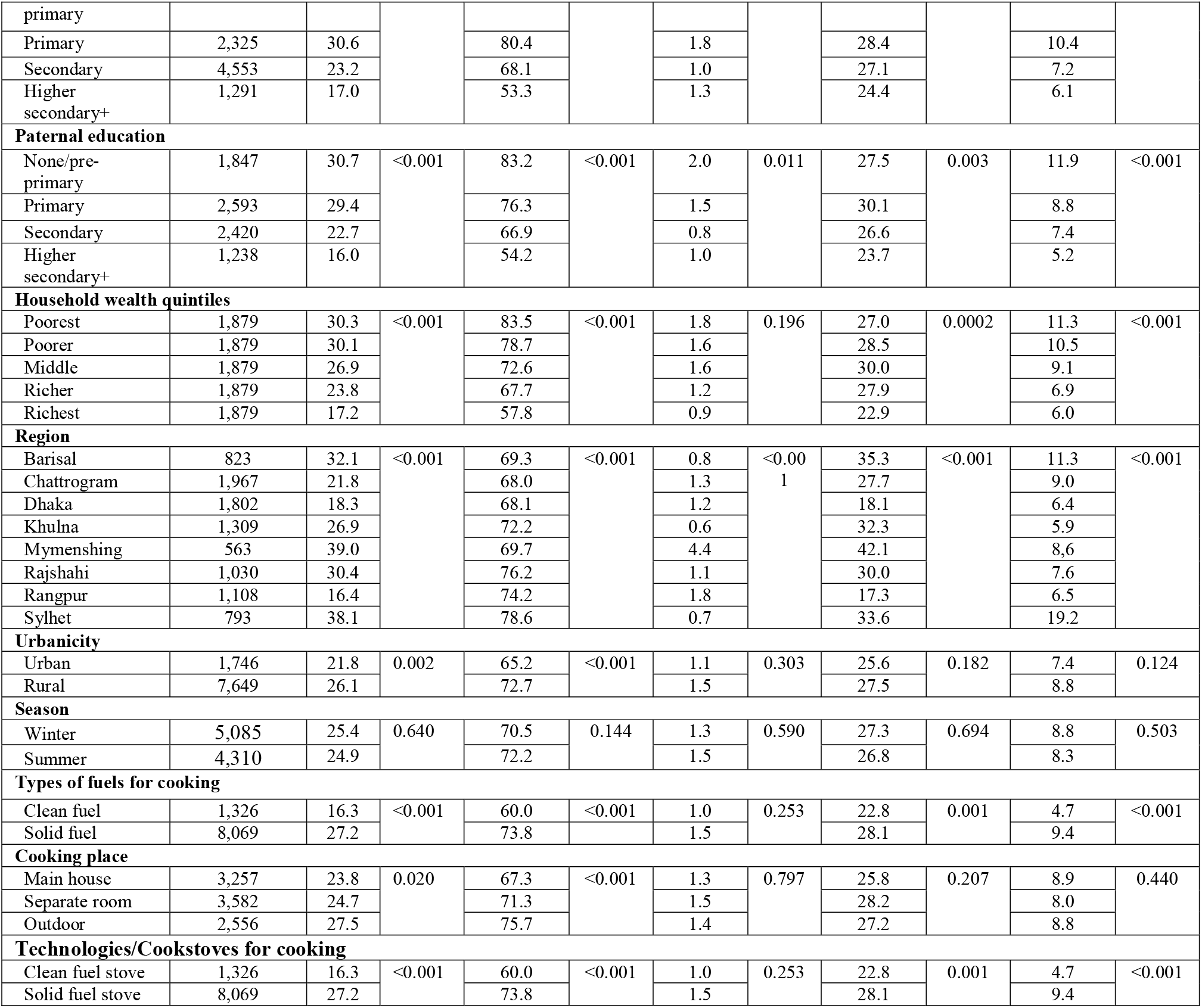
Percentage of children age 36-59 months who are developmentally not on track for indicated domains (Weighted Percentage and n=Unweighted)

All maternal, child, and household characteristics, except child size and season during the study, were significantly associated with global ECDI. Younger children (3 years old), boys, stunted children, those not receiving sufficient iodine intake, those not ever attending an ECDEP, not receiving or poor caregivers’ support, with parents below primary education, living in rural areas and poorest/poorer households and using solid fuels for cooking were more likely to be not developmentally on track in ECDI. Figure 2 shows that the highest percentages of children were not developmentally on track in global ECDI in Mymenshing (PR of ECDI 39.1) and Sylhet divisions (PR of ECDI 38.1).

**Figure 1:**
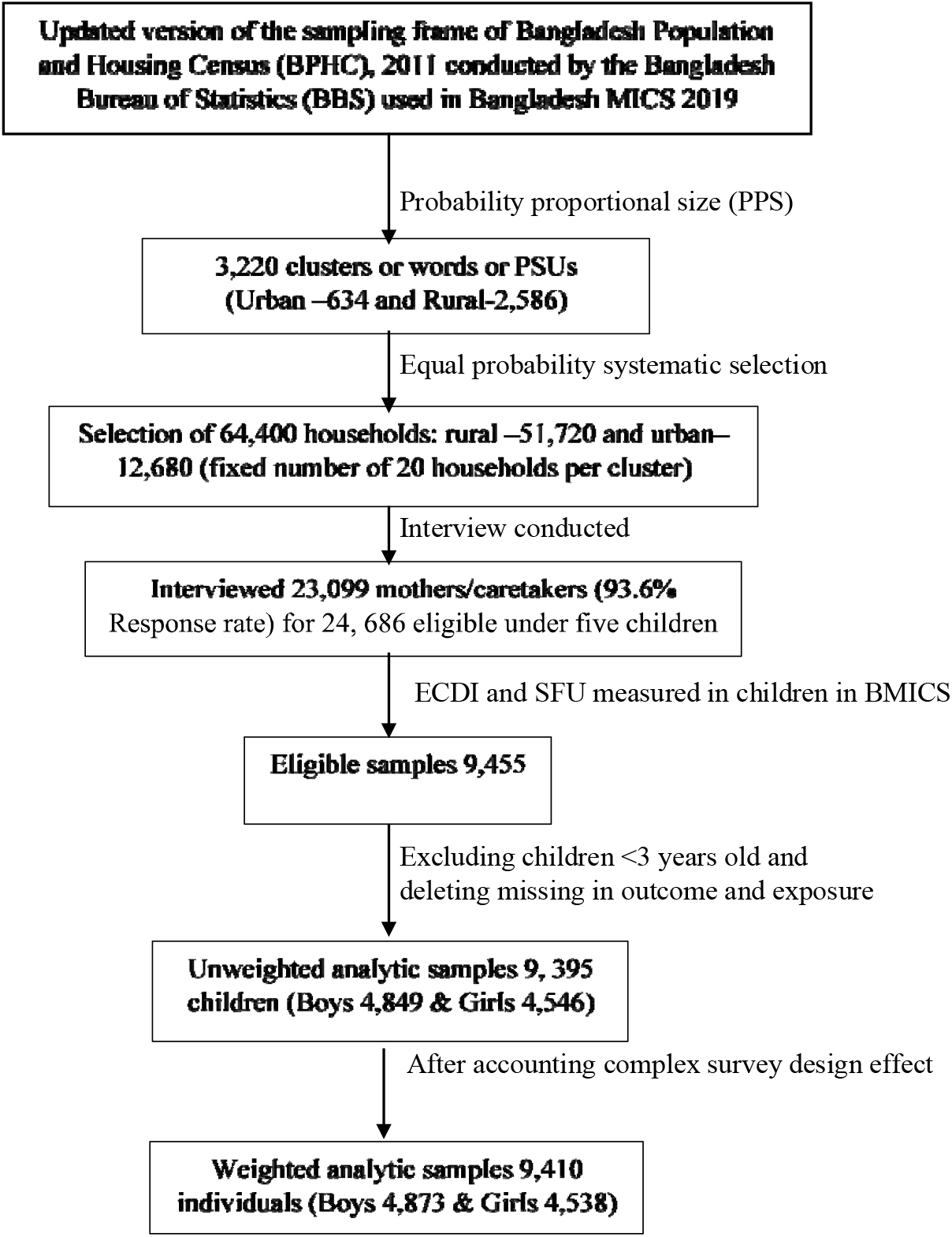
Schematic diagram of the analytic study sample.

**Figure 2:**
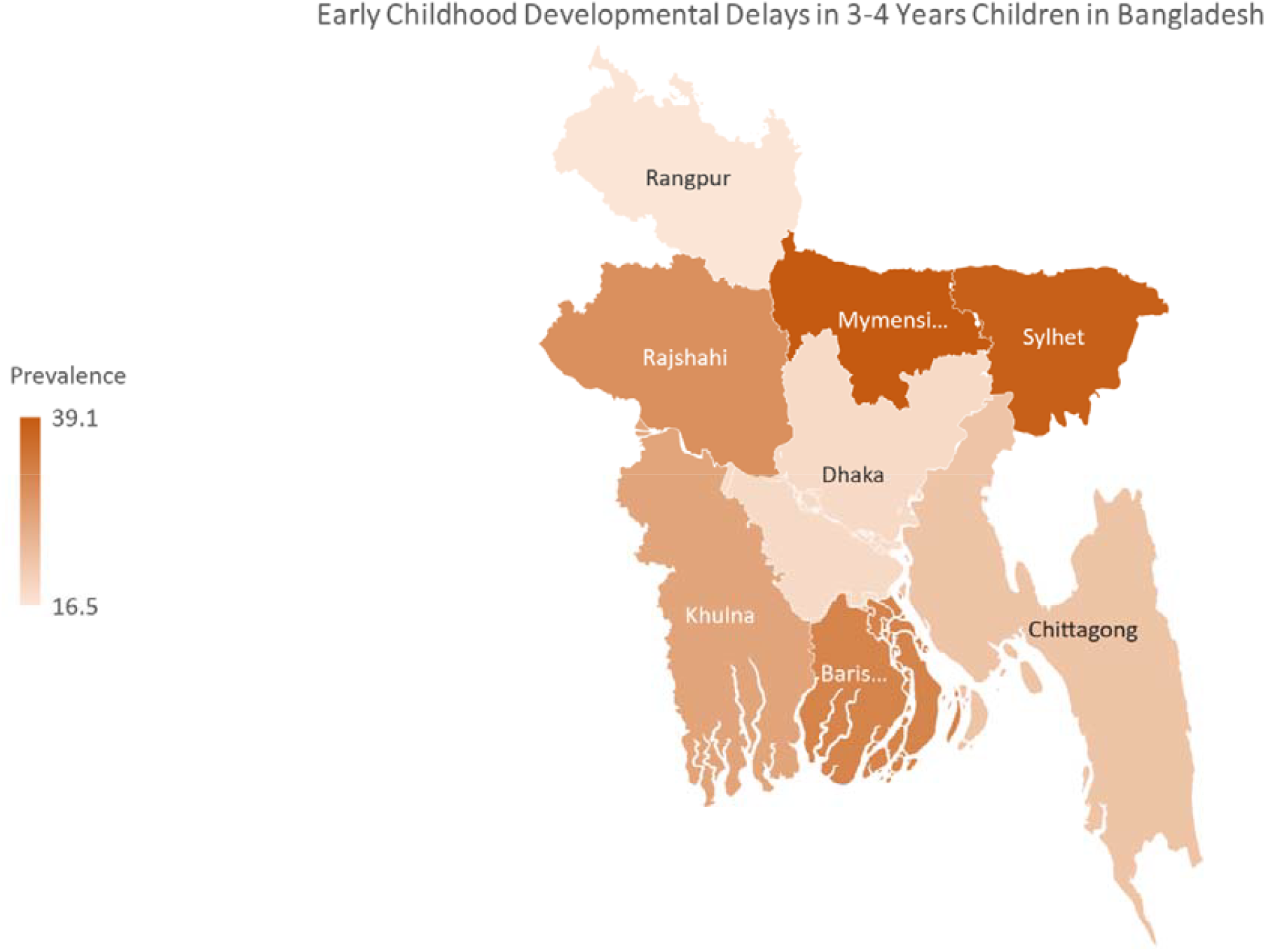
Regional variation in early childhood developmental delays.

On subdomains, all maternal, household, and children characteristics except for child size and seasonality were associated with literacy-numeracy. Only caregivers’ sufficient support, parental education, wealth quintiles, and region were associated with physical development. Regarding socio-emotional development, children’s age, sex, paternal education, wealth quintiles, region, type of cooking fuels, and cookstove technologies were associated with this domain. Finally, learning was associated with children’s age, stunting, ever attended ECDEP, parental education, wealth quintiles, region, and type of cooking fuels and cookstoves used.

### 3.2. Multivariable associations between SFU and ECD outcomes by child sex and urbanicity

**Figure 3-4** shows the prevalence ratios of the associations between ECD outcomes and SFU by sex of the children and urbanicity after adjusting for potential confounders. Children exposed to SFU were 1.47 times more likely not to be developmentally on track in global ECDI (95% CI: 1.25, 1.73; p<0.001). Similar associations were observed on regard to socio-emotional development (PR: 1.17; 95% CI: 1.01, 1.36; p=0.035) and learning-cognitive development (PR: 1.90; 95% CI: 1.39, 2.60; p<0.001) with significant differences for each outcome. However, literacy-numeracy (PR: 1.04; 95% CI: 0.95, 1.13) and physical development (PR: 0.71, 95% CI: 0.32, 1.60) did not show any significant associations with SFU.

**Figure 3:**
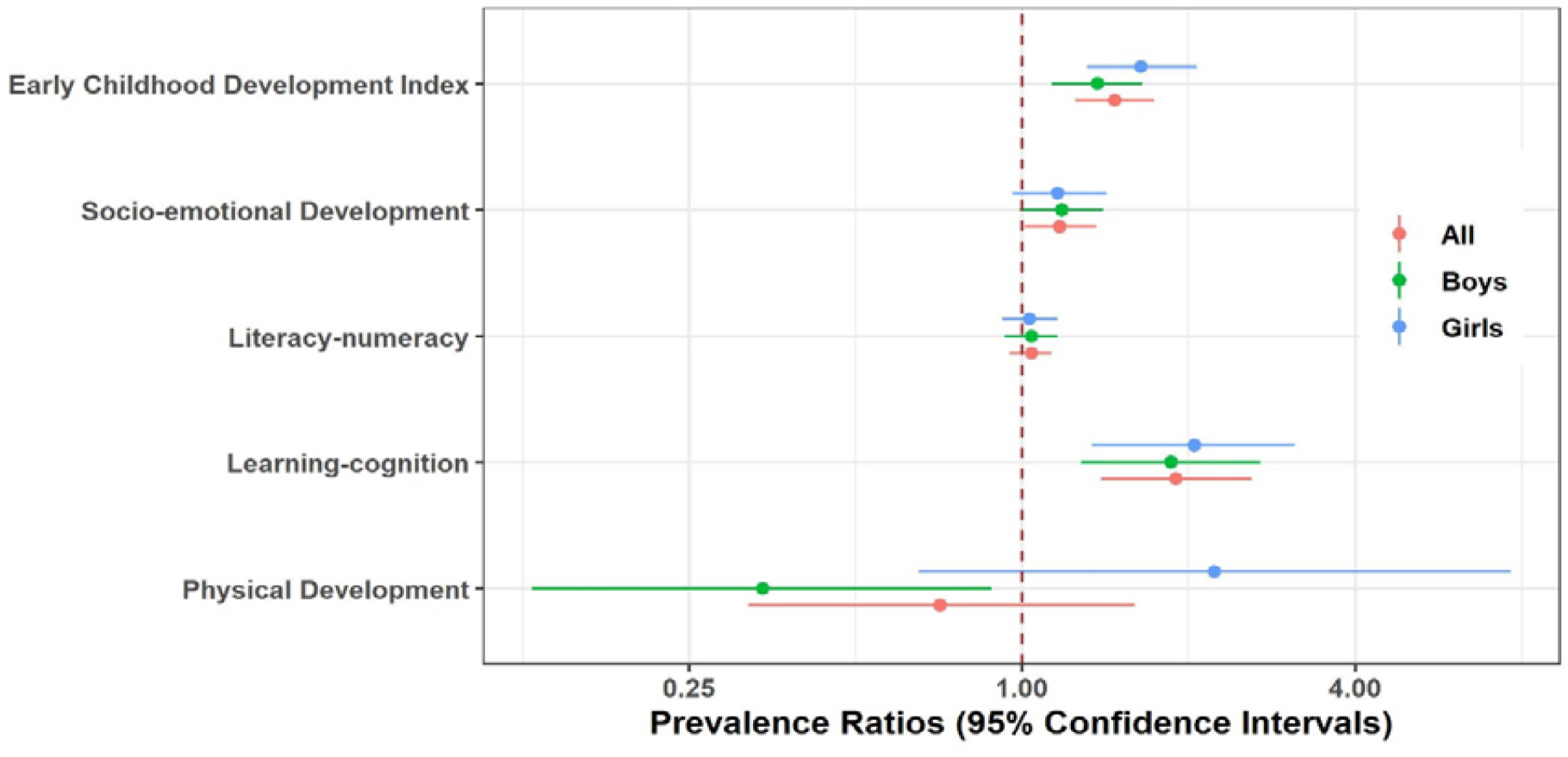
Association between SFU and ECD outcomes stratified by sex of the children in Bangladesh. *Models were adjusted for child age, maternal age, maternal education, household wealth quintiles, iodine, stunting, attendance to ECDEP, cooking place, urbanicity, and season (Models in table are included as supplemental files 1,3,5,7,9).

**Figure 4:**
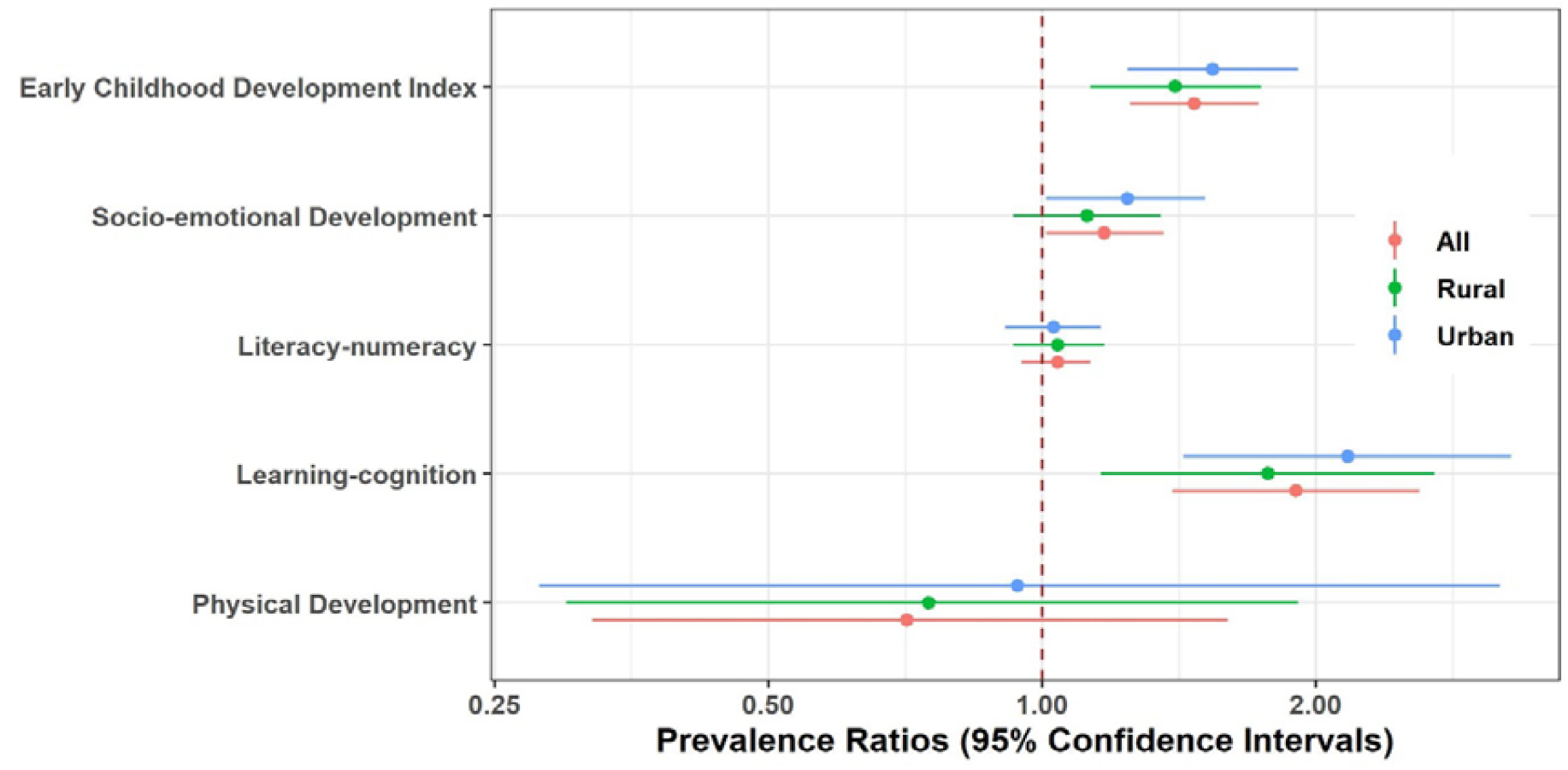
Association between SFU and ECD outcomes stratified by urbanicity of the children in Bangladesh. * Models were adjusted for child sex, child age, maternal age, maternal education, household wealth quintiles, iodine, stunting, attendance to ECDEP, cooking place, and season (Models in table are included as supplemental files 2,4,6,8,10).

When exploring the effect modification by the sex of the children (Figure 3), we did not observe significant effect modification, and results were similar (p-difference= 0.210) between boys (PR: 1.37; 95% CI: 1.13, 1.65) and girls (PR: 1.64; 95% CI: 1.31, 2.07) for ECDI. Similar findings were observed for ECD sub-domains (p-difference > 0.8 for all).

When exploring the effect modification by urbanicity (Figure 4), we did not observe significant effect modification and were not significantly different (p-difference=0.450) between urban (PR: 1.54; 95% CI: 1.24, 1.91) and rural (PR: 1.40; 95% CI: 1.13, 1.74) children for global ECDI outcome. Similar effect results were observed for ECDI sub-domains (p-difference>0.2 for all).

## 4. Discussion

Our study demonstrates that Bangladeshi children aged 36-59 months exposed to SFU were more likely to be not developmentally on track compared to unexposed children. The results show that children exposed to SFU were more likely to have developmental delays, specifically in literacy-numeracy, socio-emotional development, and learning-cognition. We used nationally representative MICS data collected by UNICEF, in which SFU was considered as a proxy for HAP (37–40). These results are in line with a study conducted in Sri Lanka, in which the neurotoxic impact of HAP on child’s cognitive scores was observed (32). Similar experiences were observed in many other Asian countries in which SFU is highly prevalent (41–43). In these settings, it is also common for mothers to cook in the same room, where the majority of the children’s daily activities take place, as they mostly do not have a separate cooking place (10). Hence, the complex mixture of pollutants emitted from SFU increases the risk of developmental delays in younger children as they have the potential for damaging the central nervous system and affect brain development (14,25,44,45). The findings also showed that SFU was significantly associated with specific developmental sub-domains in both boys and girls.

### 4.1. SFU and ECD outcomes stratified by sex of the child

Unlike other studies (23) in which differences between boys and girls were found in Ghana, our study did not confirm statistically significant effect modification of sex. One reason could be methodological and another theoretical. The study from Ghana did not use seasonality and place where the cooking occurs, but in our study, we have adjusted the place of cooking in the households and seasonality in the multilevel models. Theoretically, cultural practices between Ghana and Bangladesh could be different as well. For instance, boys and girls may spend about the same time in outdoor activities in the South-Asian countries. Moreover, a previous study showed that sex-dependent effects on neuropsychological development at 4–6 years of age, with a greater vulnerability in boys, specifically in domains related to memory, verbal and general cognition (38). This study also found that other confounders such as parents’ education, child age, and wealth index are significantly associated with global ECDI.

As such, this study contributes to setting specific targets and tracking progress towards meeting SDGs of 4.2: “By 2030, ensure that all girls and boys have access to quality early childhood development, care, and pre-primary education so that we can save the brain of our next generation (5,6).” Thus, it is pressing to design interventions and implementation strategies for lessening the HAP exposure from SFU and cooking places and utilizing low toxic supported technologies for cooking and improving cooking conditions to reduce the risk of younger children not being developmentally on track in LMICs, including Bangladesh.

### 4.2. SFU and ECD outcomes stratified by urbanicity

We observe that urban children exposed to SFU were more likely to be delayed in ECDI and its sub-domains of socio-emotional development and learning-cognition compared to rural children, but these differences were not statistically significant. However, rural-urban differences in ECD outcomes may be because children in rural areas have more mobility and spend more time playing outdoors with other children of joint families, relatives, and neighbors. *Heissler* also showed that it is common for rural children to grow-up with other family members, including grandparents, uncles, and same-aged children in Bangladesh (46). However, there is less opportunity to play with other children and spend time with other family members and relatives who can provide more nurturing (46). Interestingly, urban children showed more learning-cognition delays compared to rural children, maybe because most urban children stay home and have less opportunity for outdoor activities. Simultaneously, it is also important to investigate children’s outdoor playing time since this may also limit the effects of indoor pollution areas relatively free of external contamination, especially in rural areas as urban areas in Bangladesh experience high outdoor air pollution levels.

### 4.3. Strengths and limitations

Despite the interesting findings of this study, there are some strengths and weaknesses. This study’s primary strength is a large sample size with a representative sample of children from the whole country. Another strength is that we adjusted for potential confounders-socioeconomic and demographic variables, including child sex and age, maternal age and education, household wealth and attendance to ECDEP, and lifestyle variables are known to influence child development. However, this study has some limitations. First, our primary SFU exposure was constructed only using cooking fuel types with no direct measurements of household air pollutants’ levels. In Bangladesh, many households in rural areas use animal dung or agricultural crop, which are more toxic for health and development (45). It is also essential to consider other sources of household pollutants co-exist. In this regard, it would be beneficial if MICS would collect other types of fuels’ information since this survey only provides information on SFU.

Last, the cross-sectional and observational nature of the study may not allow us to identify with more precision how SFU directly affects ECDI and to establish any specific causal link at this time. Further longitudinal studies are needed to confirm more precise causal effects of SFU on the ECD outcomes and better understand the impacts of clean cookstove interventions on child developmental milestones. However, it has been shown that estimates of the prevalence of household SFU from multi-stage probability sampling surveys can be made with more confidence than estimates of actual pollutant exposures (7). In this study, there might have recall biases on each component of ECDI. However, we have adjusted potential confounders in our analyses to minimize its effects on estimates and data collected from the whole country, which increases our findings’ generalizability. Despite these limitations, the lack of studies investigating the effects of SFU on child neurodevelopment in LMICs makes this study valuable and a starting point to further investigations with a better assessment of exposure to HAP, including information on stove type, cooking/heating practices, and time-activity patterns.

## 5. Conclusion

This study showed that children exposed to SFU might have significant developmental delays compared to unexposed children, regardless of the child’s sex or urbanicity. Hence, we should implement effective interventions to reduce SFU exposure for ensuring better early child development in LMICs, including Bangladesh. Specifically, it is urgent to design programs to reduce HAP and its health effects, as it is a double burden for most disadvantaged kids struggling to reach their full developmental potentials (47,48). Through an effective intervention, public policies should promote a better early life environment for our younger children to meet their developmental milestones. Hence, the policymakers must take initiatives and policies regarding clean cookstoves to reduce SFU-related developmental delays in young children. Therefore, young children can reach their full developmental potential; such a program will promote and nurture the healthy and productive early brain development of our next generation.

## Data Availability

Detailed information on the survey is available at http://mics.unicef.org/

http://mics.unicef.org/

## Acknowledgments

The authors are grateful to BBS and UNICEF for granting access to the Bangladesh MICS 2019 Data.

## Ethical Approval

The technical committee of the Government of Bangladesh lead by BBS approved the MICS 2019 survey protocol. Informed consent was taken from each participant, and adult consent was taken for the child’s assent before the enrollment. We obtained approval from UNICEF in March 2020 to use this publicly available data of the MICS online archive’s de-identified data.

